# Post COVID-19 in children, adolescents, and adults: results of a matched cohort study including more than 150,000 individuals with COVID-19

**DOI:** 10.1101/2021.10.21.21265133

**Authors:** Martin Roessler, Falko Tesch, Manuel Batram, Josephine Jacob, Friedrich Loser, Oliver Weidinger, Danny Wende, Annika Vivirito, Nicole Toepfner, Martin Seifert, Oliver Nagel, Christina König, Roland Jucknewitz, Jakob Peter Armann, Reinhard Berner, Marina Treskova-Schwarzbach, Dagmar Hertle, Stefan Scholz, Stefan Stern, Pedro Ballesteros, Stefan Baßler, Barbara Bertele, Uwe Repschläger, Nico Richter, Cordula Riederer, Franziska Sobik, Anja Schramm, Claudia Schulte, Lothar Wieler, Jochen Walker, Christa Scheidt-Nave, Jochen Schmitt

**Author notes:** Corresponding author: Martin Roessler, Center for Evidence-Based Healthcare (ZEGV), University Hospital Carl Gustav Carus and Carl Gustav Carus Faculty of Medicine, TU Dresden, Fetscherstr. 74, 01307 Dresden, Germany.

## Abstract

**Background:** Long-term health sequelae of the coronavirus disease 2019 (COVID-19) are a major public health concern. However, evidence on post-acute COVID-19 syndrome (post COVID-19) is still limited, particularly for children and adolescents. Utilizing comprehensive healthcare data on more than 45 percent of the German population from January 2019 through December 2020, we investigated post COVID-19 in children/adolescents and adults.

**Methods:** From a total of 38 million individuals, we identified all patients with laboratory confirmed diagnosis of COVID-19 through June 30, 2020. A control cohort was assigned using 1:5 exact matching on age, sex, and propensity score matching on prevalent medical conditions. COVID-19 and control cohorts were followed for incident morbidity outcomes documented at least three months after the date of COVID-19 diagnosis, which was used as the index date for both groups. Overall, 96 pre-defined outcomes were aggregated into 13 diagnosis/symptom complexes and three domains (physical health, mental health, physical/mental overlap domain). We used Poisson regression to estimate incidence rate ratios (IRRs) with 95%-confidence intervals (95%-CI).

**Results:** The study population included 157,134 individuals (11,950 children/adolescents and 145,184 adults) with confirmed COVID-19. COVID-19 and control cohort were well-balanced regarding covariates. For all health outcomes combined, incidence rates (IRs) in the COVID-19 cohort were significantly higher than those in the control cohort in both children/adolescents (IRR=1.30, 95%-CI=[1.25-1.35], IR COVID-19=436.91, IR Control=335.98) and adults (IRR=1.33, 95%-CI=[1.31-1.34], IR COVID-19=615.82, IR Control=464.15). The relative magnitude of increased documented morbidity was similar for the physical, mental, and physical/mental overlap domain. In the COVID-19 cohort, incidence rates were significantly higher in all 13 diagnosis/symptom complexes in adults and in ten diagnosis/symptom complexes in children/adolescents. IRR estimates were similar for the age groups 0-11 and 12-17. Incidence rates in children/adolescents were consistently lower than those in adults. Among the specific outcomes with the highest IRR and an incidence rate of at least 1/100 person-years in the COVID-19 cohort in children and adolescents were malaise/fatigue/exhaustion (IRR=2.28, 95%-CI=[1.71-3.06], IR COVID-19=12.58, IR Control=5.51), cough (IRR=1.74, 95%-CI=[1.48-2.04], IR COVID-19=36.56, IR Control=21.06), and throat/chest pain (IRR=1.72, 95%-CI=[1.39-2.12], IR COVID-19=20.01, IR Control=11.66). In adults, these included dysgeusia (IRR=6.69, 95%-CI=[5.88-7.60], IR COVID-19=12.42, IR Control=1.86), fever (IRR=3.33, 95%-CI=[3.01-3.68], IR COVID-19=11.53, IR Control=3.46), and dyspnea (IRR=2.88, 95%-CI=[2.74-3.02], IR COVID-19=43.91, IR Control=15.27).

**Conclusions:** This large, matched cohort study indicates substantial new-onset post COVID-19 morbidity in pediatric and adult populations based on routine health care documentation. Further investigation is required to assess the persistence and long-term health impact of post COVID-19 conditions, especially in children and adolescents.

## Background

There is mounting evidence that a yet unknown proportion of persons suffers from long-term complications after severe acute respiratory syndrome coronavirus type 2 (SARS-CoV-2) infection. Early in the pandemic, patients started to share their experiences in the social media on what they referred to as “Long-haul COVID” or “Long COVID.” They reported a wide variety of somatic and mental health issues that were either persisting, recurring, or newly occurring beyond the 4-week phase of acute coronavirus disease 2019 (COVID-19) and even mild or asymptomatic SARS-CoV-2 infection [1]. Health organizations and medical societies at the national and international level have taken efforts to systemize observations from an increasing body of observational research studies, in order to provide a pragmatic case definition for this new health phenomenon. Most recently, a WHO working group including patients and a multidisciplinary team of medical experts proposed a preliminary clinical case definition using the term “post COVID-19 condition.” Based on a structured consensus process, this definition refers to a broad spectrum of otherwise unexplained health conditions that are present three months after the onset of symptoms or date of SARS-CoV-2 infection and last for at least two months [2]. Notably, it is still uncertain whether the definition applies to adults as well as children and adolescents due to the paucity of available data among younger age groups [2].

Until now, few clinical and epidemiological studies on long-term health consequences of SARS-CoV-2 infection have followed patients for at least three months after SARS-CoV-2 infection. The vast majority of these studies focused solely on adults [3–7]. Results of these studies are heterogenous, due to large methodological differences with regard to study design, data sources, data collection modes, and the definition of health outcomes [3,4,8–10]. Clinical studies of patients hospitalized for COVID-19 have consistently reported that high proportions of patients continue to suffer from one or more health complaints for months after discharge. Population-based studies, also including persons with a mild course of COVID-19, have reported much lower proportions of persons affected with symptoms beyond three months after confirmed or suspected SARS-CoV-2 infection. Most of these studies observed that symptoms decline over time. Symptoms frequently observed include fatigue, weakness, muscle pain, sleep disorders, depressive symptoms, dizziness, lack of memory and concentration, and shortness of breath. This has been observed mainly among adults [8,9,11], whereas evidence on pediatric populations is still very limited [10,12,13].

A recently published review [12] identified only 14 studies on persistence of symptoms following COVID-19 in children and adolescents. The authors concluded that almost all existing studies have major limitations such as lack of a clear case definition, absence of control groups, low follow-up rates, small sample sizes resulting in limited statistical power, reliance on self- or parent-reported symptoms without clinical assessment, and non-response and other forms of selection and information biases [12]. The current uncertainty regarding the incidence and health impact of post COVID-19 among children and adolescents has hampered health policy decisions regarding social and medical preventive measures for this population group.

Against that background, we investigated documented long-term morbidity of COVID-19 based on routine data from multiple German statutory health insurances. We hypothesized that SARS-CoV-2 infections induce higher morbidity three month after first diagnosis of COVID-19 or later in both children/adolescents and adults compared to controls without previous COVID-19. We expected this higher morbidity to be reflected in more intensive utilization of healthcare services and, hence, diagnoses documented by physicians. To account for the heterogeneity of potential long-term sequelae of COVID-19 reported in the literature [7,9], we took a sensitive empirical approach and explored relationships between COVID-19 and a large set of outcomes covering multiple organ systems and diagnosis/symptom complexes.

## Methods

### Study design

The Post-COVID-19 Monitoring in Routine Health Insurance Data (POINTED) program was established in December 2020 as part of the German Network University Medicine project egePan Unimed, which focuses on the development, evaluation, and implementation of evidence-based pandemic management. The overarching aim of the POINTED program is to rapidly generate new evidence on prioritized questions concerning pandemic response with substantial public health impact by the use of healthcare data. The POINTED consortium is coordinated by the Center for Evidence-Based Healthcare (ZEGV) at the TU Dresden and consists of large German statutory health insurances, health services research institutes (ZEGV and InGef - Institute for Applied Health Research Berlin), the Robert Koch Institute (RKI), and clinical experts.

As part of the POINTED program, we designed a matched cohort study based on routine health insurance data. The exposure cohort included individuals with a confirmed first COVID-19 diagnosis as of June 30, 2020. In addition to exact matching by age and sex, we applied propensity score matching based on data from 2019 providing information on prevalent health conditions. The control cohort included individuals without COVID-19 diagnosis in 2020. The entire observation period thus was from January 1st, 2019 to December 31st, 2020. Individuals included in the COVID-19 or control cohort were followed over at least three months for a set of pre-defined incident health conditions. The date of the COVID-19 diagnosis among individuals with COVID-19 was used as the index date for individuals in the COVID-19 cohort as well as for matched controls. Assigning identical index dates to matched COVID-19 and control persons offers the advantages that potential follow-up times are identical and that the timing of non-pharmaceutical interventions (e.g. lockdowns) is equally captured in both cohorts.

The study was registered at ClinicalTrials.gov (NCT number: NCT05074953). The competent authority of the Federal State of Saxony, Germany approved the study protocol and declared waiver of informed consent (reference number: 31-5221.40-15/68). The study was approved by the ethics committee of the TU Dresden (approval number: BO-EK (COVID)-482102021) and adheres to the Declaration of Helsinki and all relevant administrative and legal regulations.

### Data

We used routine data from six German statutory health insurances: AOK Bayern - Die Gesundheitskasse, AOK PLUS (analyzed by ZEGV), BARMER, BKKen (analyzed by InGef), DAK Gesundheit (analyzed by Vandage GmbH), and Techniker Krankenkasse. In total, these data cover approximately 38 million persons, which corresponds to 51.8% of all persons insured with German statutory health insurances and 45.7% of the total German population. In addition to sociodemographic characteristics (age and sex) and vital status (via the date of death), the data include comprehensive information on healthcare utilization in outpatient and inpatient sectors. The data include diagnoses (according to the International Statistical Classification of Diseases and Related Health Problems - German Modification, ICD-10-GM), procedures (according to the “Operationen-und Prozedurenschluessel,” OPS; German modification of the International Classification of Procedures in Medicine, ICPM), information on outpatient medical services (according to “Einheitlicher Bewertungsmassstab,” EBM), and prescribed medications (according to the German Anatomical Therapeutic Chemical (ATC) Classification).

### Cohorts

The full COVID-19 cohort included individuals with documented COVID-19 diagnosis with confirmed laboratory virus detection (ICD-10-GM: U07.1!) in 2020. The control cohort included individuals *without* COVID-19 diagnosis *regardless of laboratory virus detection* (ICD-10-GM: U07.1! or U07.2!) in 2020. We excluded individuals with COVID-19 diagnosis without laboratory virus detection (ICD-10-GM: U07.2!) from both groups to avoid distortions due to misclassification. We further excluded persons who were not continuously insured with the respective health insurance between 2019-01-01/birth and 2020-12-31/death because relevant outcomes and prevalent health conditions may not have been documented in our data.

### Follow-up

Currently, there is no agreed-upon standard case definition of post COVID-19. We therefore followed the NICE guideline on long COVID [14] and the working clinical case definition of post COVID-19 condition proposed by the WHO [2], and considered an individual to enter the post COVID-19 phase three months after diagnosis of COVID-19. Due to the characteristics of the German healthcare billing system, outpatient diagnoses in our data can be assigned reliably to a specific quarter of the year. Accordingly, we considered a diagnosis to have been made in the post COVID-19 phase if it was documented in the second quarter after index date or later. This operationalization ensures a time distance of at least three months between date of COVID-19 diagnosis and post COVID-19 outcome incidence. Consequently, we excluded individuals with first COVID-19 diagnosis later than June 30st, 2020 from post COVID-19 analyses due to insufficient follow-up time. We also excluded individuals who died before reaching the required minimum follow-up time.

### Health outcomes

Based on published literature and clinical expertise in the author team, we defined 96 potential post COVID-19 health outcomes. These outcomes constitute new-onset morbidity documented by a physician or psychotherapist within the statutory healthcare system. Operationalization of these outcomes was based on inpatient and outpatient diagnoses according to ICD-10-GM and the guidelines good practice secondary data analysis (GPS) of the German Society for Epidemiology (DGEpi) [15]. In addition, we combined these outcomes into 13 diagnosis/symptom complexes and three outcome domains (physical health, mental health, physical/mental overlap domain). An overview of the outcomes and their grouping is provided in the supplementary material S1. Aggregation of outcomes into groups offers the advantage of higher statistical power, particularly in the case of post COVID-19, which is considered to include multiple rare symptoms and diagnoses [7,9,14]. However, aggregation may also obscure relevant heterogeneity in outcomes. Hence, our analysis considered both aggregate outcome groups and specific outcomes.

For each individual, we classified an outcome as prevalent if it was diagnosed at least once in the four quarters preceding the index date. Given the prevalence status, we defined post COVID-19 outcome incidence as a diagnosis of a non-prevalent outcome in the post COVID-19 phase.

### Covariates

For each individual, we used information on prevalent health conditions in the four quarters preceding the index date. We selected prevalent health conditions potentially confounding the association between exposure (COVID-19) and incident health outcomes based on published evidence and clinical expertise. These included 13 prevalent medical conditions for children/adolescents and 38 prevalent medical conditions for adults (see supplementary material S2). In line with previous studies [6,7], we also considered age and sex as well as the severity of COVID-19 as a covariate with potential influence on post COVID-19 by distinguishing between 1) individuals with outpatient diagnoses of COVID-19 only, 2) individuals with at least one hospital visit with COVID-19 diagnosis, 3) individuals with intensive care and/or ventilation (ICU) with COVID-19 diagnosis.

## Statistical methods

### Matching

To minimize differences between COVID-19 and control cohort in terms of covariates that may confound relationships between outcomes and exposure, we applied 1:5-matching with replacement [16]. For each individual in the COVID-19 cohort, we selected five control persons with identical age (in years) and sex. We chose exact matching on these characteristics to facilitate stratified analysis, e.g. for different age groups. In addition, we accounted for the presence of the medical conditions mentioned above by propensity score matching. Estimation of the propensity score was based on logistic regression including all insured persons. Given different sets of medical conditions considered as covariates, we estimated separate regression models for children/adolescents and adults.

### Modeling

We estimated differences between COVID-19 and control cohort regarding incidence rates (IRs) of outcomes per 1,000 person-years using Poisson regression [17]. Poisson regression facilitates adjustment for differences in individual-specific times at risk (time between index date and end of observation period or death) due to inclusion of these times as offset in the model. Based on the results of Poisson regressions, we derived incidence rate ratios (IRRs) with 95%-confidence intervals (95%-CI) to characterize relative incidence in COVID-19 and control cohort. Statistical significance of estimated IRRs was assessed using significance levels of 0.05 and 0.01.

We excluded individuals from the analysis of post COVID-19 incidence if the considered outcome was prevalent before the index date. To maintain balance of cohorts regarding covariates, we excluded a complete matched group of COVID-19 and control persons if the outcome was prevalent for the individual with COVID-19 or all of his/her matched control persons. For estimation, we weighted data from individuals in the control cohort with the inverse number of persons remaining in the respective matched group (i.e. weights between 1/5 and 1) to ensure that total weights in the control cohort add up to the number of persons in the COVID-19 cohort.

### Evidence synthesis

Since pooling of individual-level data was not possible due to data protection restrictions, the six health insurance datasets were analyzed separately by authorized institutes or the healthcare research department within the respective health insurance. To synthesize evidence across datasets, we used the fact that Poisson regression models can be estimated based on individual-level or aggregate data with identical point estimates [18]. Each authorized institute calculated the required aggregate statistics and provided them to ZEGV, where regressions based on combined aggregate data were performed. While point estimates are identical, variance estimates based on aggregate matched data tend to be larger than those based on matched individual-level data. Hence, our results are conservative in terms of statistical significance.

## Results

### Descriptive statistics

Our full COVID-19 cohort included 678,965 individuals (57,763 children/adolescents and 621,202 adults) with COVID-19 diagnosis in all four quarters of 2020 (Table 1). 157,134 (23.1%) of these individuals (11,950 children/adolescents and 145,184 adults) received their first COVID-19 diagnosis before July 2020 and, thus, were included in the post COVID-19 sample as they could be observed at least three months after first diagnosis. Prevalence of medical conditions selected as covariates was generally lower in children/adolescents than in adults. While our post COVID-19 sample included 8,407 (5.8%) hospitalized adults and 3,075 (2.1%) adults with intensive care and/or ventilation, smaller proportions of children/adolescents were hospitalized with COVID-19 (n=117; 1.0%) and received intensive care and/or ventilation (n=51; 0.4%). The distributions of covariates in COVID-19 and the matched control cohort were similar for both children/adolescents and adults, which indicated successful balancing (see appendix, Tables A1 and A2 for matching results).

**Table 1:** Characteristics of individuals with COVID-19 included in full COVID-19 and post COVID-19 sample

| Variable | Category | n (full COVID-19 sample) | Percent (full COVID-19 sample) | n (post COVID-19 sample) | Percent (post COVID-19 sample) |
| --- | --- | --- | --- | --- | --- |
| Children/adolescents |  | 57,763 | 100% | 11,950 | 100% |
| Age | 0-11 | 31,345 | 54.3% | 8,032 | 67.2% |
|  | 12-17 | 26,418 | 45.7% | 3,918 | 32.8% |
| Sex | female | 28,120 | 48.7% | 5,745 | 48.1% |
|  | male | 29,643 | 51.3% | 6,205 | 51.9% |
| Asthma |  | 595 | 1.0% | 128 | 1.1% |
| Bronchopulmonary dysplasia |  | 30 | 0.1% | 7 | 0.1% |
| Cancer |  | 130 | 0.2% | 28 | 0.2% |
| Congenital heart disease |  | 868 | 1.5% | 253 | 2.1% |
| Diabetes: type I with insulin |  | 173 | 0.3% | 34 | 0.3% |
| Dialysis |  | 9 | 0.0% | <5 |  |
| Down syndrome |  | 70 | 0.1% | 20 | 0.2% |
| Epilepsy |  | 501 | 0.9% | 107 | 0.9% |
| Immunosuppressive disease |  | 474 | 0.8% | 113 | 0.9% |
| Immunosuppressive therapy |  | 110 | 0.2% | 33 | 0.3% |
| Obesity |  | 79 | 0.1% | 15 | 0.1% |
| Primary immunodeficiency |  | 263 | 0.5% | 62 | 0.5% |
| Psychomotor deficit |  | 1,577 | 2.7% | 381 | 3.2% |
| Severity of COVID-19 | outpatient | 56,827 | 98.4% | 11,782 | 98.6% |
|  | hospital | 723 | 1.3% | 117 | 1.0% |
|  | ICU | 213 | 0.4% | 51 | 0.4% |
| Adults |  | 621,202 | 100% | 145,184 | 100% |
| Age | 18-24 | 60,425 | 9.7% | 12,815 | 8.8% |
|  | 25-39 | 150,971 | 24.3% | 36,565 | 25.2% |
|  | 40-49 | 100,624 | 16.2% | 24,823 | 17.1% |
|  | 50-54 | 63,011 | 10.1% | 16,291 | 11.2% |
|  | 55-59 | 63,260 | 10.2% | 16,332 | 11.2% |
|  | 60-64 | 45,132 | 7.3% | 11,599 | 8.0% |
|  | 65-69 | 25,082 | 4.0% | 6,035 | 4.2% |
|  | 70-74 | 20,886 | 3.4% | 4,700 | 3.2% |
|  | 75-79 | 23,047 | 3.7% | 4,586 | 3.2% |
|  | 80-plus | 68,764 | 11.1% | 11,438 | 7.9% |
| Sex | female | 358,777 | 57.8% | 87,395 | 60.2% |
|  | male | 262,425 | 42.2% | 57,789 | 39.8% |
| Asthma |  | 17,694 | 2.8% | 4,533 | 3.1% |
| Autoimmune disease |  | 50,193 | 8.1% | 12,208 | 8.4% |
| Cardiac arrhythmia |  | 35,087 | 5.6% | 6,169 | 4.2% |
| Cerebrovascular disease |  | 43,354 | 7.0% | 8,521 | 5.9% |
| Chronic kidney disease |  | 48,396 | 7.8% | 8,194 | 5.6% |
| COPD or severe lung disease |  | 19,557 | 3.1% | 4,628 | 3.2% |
| Coronary heart disease |  | 39,389 | 6.3% | 7,053 | 4.9% |
| Crohn's disease |  | 2,865 | 0.5% | 777 | 0.5% |
| Dementia |  | 36,104 | 5.8% | 5,695 | 3.9% |
| Depression |  | 48,221 | 7.8% | 11,613 | 8.0% |
| Diabetes (without insulin) |  | 51,040 | 8.2% | 9,135 | 6.3% |
| Diabetes: type I with insulin |  | 5,173 | 0.8% | 1,161 | 0.8% |
| Diabetes: other with insulin |  | 23,236 | 3.7% | 4,177 | 2.9% |
| Dialysis |  | 3,310 | 0.5% | 762 | 0.5% |
| Down syndrome |  | 694 | 0.1% | 159 | 0.1% |
| Heart failure |  | 39,589 | 6.4% | 6,653 | 4.6% |
| Hemato-oncological disease with therapy |  | 1,068 | 0.2% | 241 | 0.2% |
| Hemato-oncological disease without therapy |  | 2,863 | 0.5% | 594 | 0.4% |
| Hepatitis |  | 1,860 | 0.3% | 428 | 0.3% |
| HIV |  | 914 | 0.1% | 273 | 0.2% |
| Hypertension |  | 155,603 | 25.0% | 31,371 | 21.6% |
| Immunosuppressive disease |  | 16,621 | 2.7% | 3,822 | 2.6% |
| Immunosuppressive therapy |  | 11,483 | 1.8% | 2,675 | 1.8% |
| Intellectual disabilities |  | 5,496 | 0.9% | 986 | 0.7% |
| Interstitial lung disease |  | 1,406 | 0.2% | 301 | 0.2% |
| Mental illness |  | 6,178 | 1.0% | 1,381 | 1.0% |
| Metastatic cancer with therapy |  | 2,188 | 0.4% | 432 | 0.3% |
| Metastatic cancer without therapy |  | 2,102 | 0.3% | 395 | 0.3% |
| Obesity |  | 25,112 | 4.0% | 5,306 | 3.7% |
| Organ transplant |  | 955 | 0.2% | 226 | 0.2% |
| Other neurological diseases |  | 41,306 | 6.6% | 8,329 | 5.7% |
| Primary immunodeficiency |  | 1,516 | 0.2% | 418 | 0.3% |
| Rheumatological symptoms/diagnosis |  | 21,464 | 3.5% | 4,994 | 3.4% |
| Severe lung disease |  | 946 | 0.2% | 223 | 0.2% |
| Severe or cirrhotic liver disease |  | 3,219 | 0.5% | 638 | 0.4% |
| Solid cancer with therapy |  | 4,964 | 0.8% | 1,098 | 0.8% |
| Solid cancer without therapy |  | 27,348 | 4.4% | 5,821 | 4.0% |
| Ulcerative colitis |  | 3,533 | 0.6% | 873 | 0.6% |
| Severity of COVID-19 | outpatient | 548,159 | 88.2% | 133,702 | 92.1% |
|  | hospital | 55,129 | 8.9% | 8,407 | 5.8% |
|  | ICU | 17,914 | 2.9% | 3,075 | 2.1% |

### Incidence of documented health outcome groups

Considering all outcomes combined, the incidence rate of documented health problems in the COVID-19 cohort was significantly higher than that in the control cohort (upper left panel of Figure 1). This finding holds for both children/adolescents (IRR=1.30, 95%-CI=[1.25-1.35], IR COVID-19=436.91, IR Control=335.98) and adults (IRR=1.33, 95%-CI=[1.31-1.34], IR COVID-19=615.82, IR Control=464.15). Furthermore, we found significantly higher incidence rates in the COVID-19 cohort across all considered outcome domains, i.e. physical health (children/adolescents: IRR=1.31, 95%-CI=[1.24-1.38], IR COVID-19=254.58, IR Control=194.45; adults: IRR=1.39, 95%-CI=[1.37-1.41], IR COVID-19=422.87, IR Control=304.42), mental health (children/adolescents: IRR=1.39, 95%-CI=[1.28-1.52], IR COVID-19=102.17, IR Control= 73.24; adults: IRR=1.27, 95%-CI=[1.25-1.29], IR COVID-19=215.62, IR Control=169.50), and the physical/mental overlap domain (children/adolescents: IRR=1.32, 95%-CI=[1.24-1.40], IR COVID-19=209.26, IR Control=158.71; adults: IRR=1.45, 95%-CI=[1.42-1.47], IR COVID-19=278.58, IR Control=192.59). For all 13 diagnosis/symptom complexes, incidence rates in the adult COVID-19 cohort were significantly higher than those in the adult control cohort (lower left panel of Figure 1). In children/adolescents, significantly higher incidence rates were observed for ten diagnosis/symptom complexes. Estimated IRRs ranged from 1.00 (95%-CI=[0.80-1.25]; dermatological diagnosis/symptom complex) to 1.98 (95%-CI=[1.43-2.75]; vascular/coagulation diagnosis/symptom complex) in children/adolescents and from 1.11 (95%-CI=[1.07-1.14]; gynecological/urogenital diagnosis/symptom complex) to 2.62 (95%-CI=[2.53-2.71]; pulmonary diagnosis/symptom complex) in adults. Given a 162% higher post COVID-19 incidence rate in adults, pulmonary diagnosis/symptom complex showed the most pronounced overall difference between COVID-19 and control cohort. Stratified estimations for the age groups 0-11 and 12-17 yielded similar IRRs (see supplementary Figures S1 and S2).

**Figure 1:**
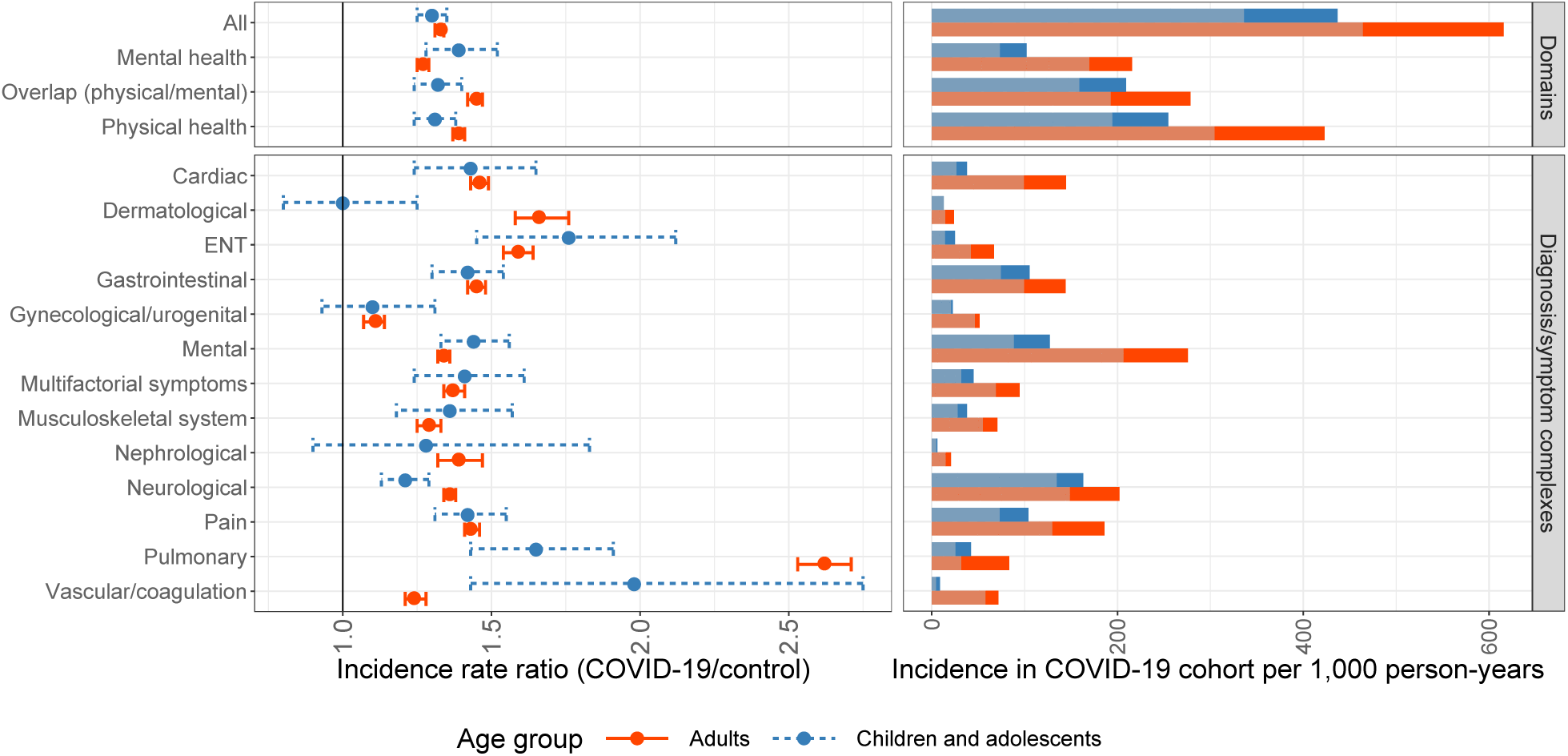
Estimated incidence rate ratios with 95%-confidence intervals and incidence rates in COVID-19 cohort for children/adolescents and adults by outcome domain and diagnosis/symptom complex Note: Incidence rates in the control cohort are shown in pale color.

Across all outcome domains and diagnosis/symptom complexes, incidence rates in the COVID-19 cohort were lower in children/adolescents than in adults (right panels of Figure 1). Regarding all outcomes combined, the incidence rate in adults with COVID-19 (IR=615.82) was 41% higher than the incidence rate in children and adolescents with COVID-19 (IR=436.91).

The full results are presented in the appendix (Tables A3 and A4).

### Incidence of documented health outcomes

To identify the most frequently documented long-term health problems among persons with COVID-19, we considered outcomes with an incidence of at least 1/100 person-years in the COVID-19 cohort. We then sorted these outcomes by IRR for children/adolescents (Table 2) and adults (Table 3). The resulting lists had five identical outcomes (cough; fever; headache; malaise/fatigue/exhaustion; throat/chest pain) across age groups. The outcomes with the highest IRR in children and adolescents were malaise/fatigue/exhaustion (IRR=2.28, 95%-CI=[1.71-3.06], IR COVID-19=12.58, IR Control=5.51), cough (IRR=1.74, 95%-CI=[1.48-2.04], IR COVID-19=36.56, IR Control=21.06), and throat/chest pain (IRR=1.72, 95%-CI=[1.39-2.12], IR COVID-19=20.01, IR Control=11.66). The outcomes with the largest IRR in adults were dysgeusia (IRR=6.69, 95%-CI=[5.88-7.60], IR COVID-19=12.42, IR Control=1.86), fever (IRR=3.33, 95%-CI=[3.01-3.68], IR COVID-19=11.53, IR Control=3.46), and dyspnea (IRR=2.88, 95%-CI=[2.74-3.02], IR COVID-19=43.91, IR Control=15.27). IRRs in adults were higher than in children/adolescents for most outcomes assessed. For all listed outcomes, estimated IRRs were statistically significant. While the unspecific diagnosis malaise/fatigue/exhaustion (ICD-10-GM: R53) was represented in the lists for both children/adolescents and adults, the chronic fatigue syndrome (ICD-10-GM: G93.3) was not. However, chronic fatigue syndrome was also coded more frequently in the COVID-19 than in the control cohort in adults (IRR=3.04, 95%-CI=[2.66-3.48], IR COVID-19=5.94, IR Control=1.95). In children, the estimated IRR was greater than 1 but not statistically significant (IRR=1.25, 95%-CI=[0.24-6.65], IR COVID-19=0.26, IR Control=0.21).

**Table 2:** 10 post COVID-19 outcomes in children/adolescents with highest IRRs and incidence of at least 1/100 person-years in the COVID-19 cohort

| Rank | Name | IRR | 95%-CI | IR COVID-19 | IR Control |
| --- | --- | --- | --- | --- | --- |
| 1 | Malaise/fatigue/exhaustion | 2.28** | [1.71-3.06] | 12.58 | 5.51 |
| 2 | Cough | 1.74** | [1.48-2.04] | 36.56 | 21.06 |
| 3 | Throat/chest pain | 1.72** | [1.39-2.12] | 20.01 | 11.66 |
| 4 | Adjustment disorder | 1.71** | [1.42-2.06] | 26.37 | 15.40 |
| 5 | Somatization disorder | 1.62** | [1.30-2.02] | 17.90 | 11.06 |
| 6 | Headache | 1.58** | [1.35-1.84] | 36.67 | 23.24 |
| 7 | Fever | 1.56** | [1.30-1.87] | 27.84 | 17.84 |
| 8 | Anxiety disorder | 1.54** | [1.23-1.92] | 16.70 | 10.87 |
| 9 | Abdominal pain | 1.45** | [1.27-1.64] | 53.94 | 37.31 |
| 10 | Depression | 1.45** | [1.12-1.87] | 12.05 | 8.32 |

Note: Significance levels: \*=5%, \*\*=1%

**Table 3:** 10 post COVID-19 outcomes in adults with highest IRRs and incidence of at least 1/100 person-years in the COVID-19 cohort

| Rank | Name | IRR | 95%-CI | IR COVID-19 | IR Control |
| --- | --- | --- | --- | --- | --- |
| 1 | Dysgeusia | 6.69** | [5.88-7.60] | 12.42 | 1.86 |
| 2 | Fever | 3.33** | [3.01-3.68] | 11.53 | 3.46 |
| 3 | Dyspnea | 2.88** | [2.74-3.02] | 43.91 | 15.27 |
| 4 | Cough | 2.80** | [2.64-2.97] | 29.95 | 10.71 |
| 5 | Respiratory insufficiency | 2.47** | [2.28-2.69] | 13.76 | 5.56 |
| 6 | Throat/chest pain | 2.20** | [2.09-2.31] | 34.57 | 15.74 |
| 7 | Hair loss | 2.02** | [1.88-2.18] | 13.96 | 6.90 |
| 8 | Malaise/fatigue/exhaustion | 1.97** | [1.89-2.06] | 42.91 | 21.74 |
| 9 | Dysphagia | 1.95** | [1.78-2.12] | 10.55 | 5.42 |
| 10 | Headache | 1.74** | [1.67-1.82] | 40.48 | 23.24 |

Note: Significance levels: \*=5%, \*\*=1%

### Incidence of documented health outcome groups by severity of COVID-19

Estimations stratified by severity indicated that IRRs for individuals with hospital visits or intensive care were higher than IRRs for individuals with outpatient diagnoses of COVID-19 only (Figure 2). This result holds for both children/adolescents and adults. However, due to the small number of children/adolescents with COVID-19-related hospital visits and intensive care and/or ventilation, precision of the corresponding IRR estimates was low as reflected in large confidence intervals.

**Figure 2:**
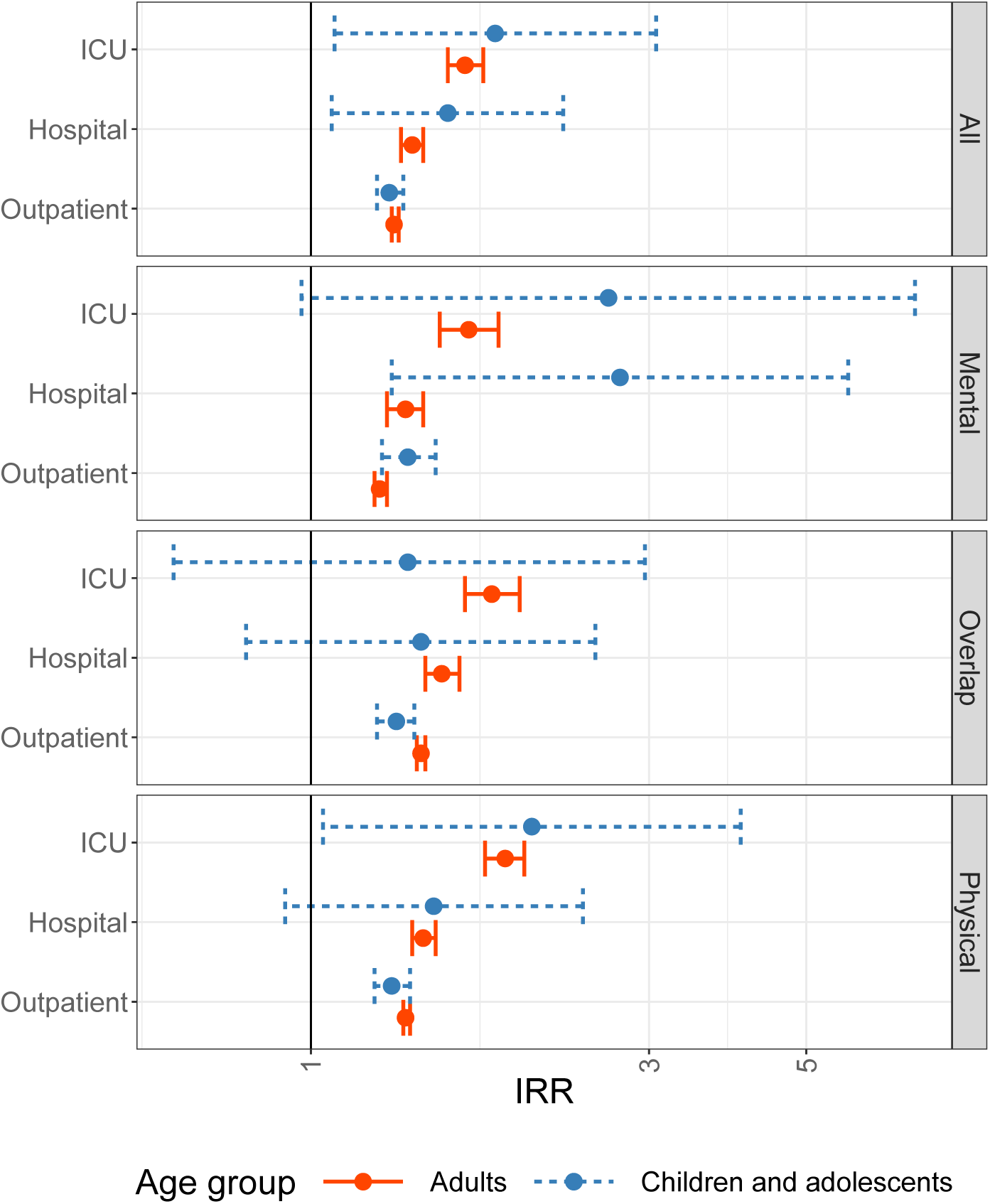
Estimated incidence rate ratios with 95%-confidence intervals in children/adolescents and adults by severity of COVID-19 and domain. Note: IRRs are shown on log-scale

## Discussion

### Summary of findings

This study makes an important contribution to the sparse empirical literature on long-term sequelae of COVID-19, particularly in children and adolescents. Thus, it addresses one of the most urgent and relevant questions at the current stage of the COVID-19 pandemic. Based on data from 157,134 individuals with confirmed COVID-19, we provide novel evidence on documented post COVID-19 morbidity in both children/adolescents and adults. Our findings indicate a higher incidence of predefined adverse health outcomes documented in routine healthcare data not only among adults but also in children and adolescents with COVID-19 compared to matched controls. This was true across all outcome domains (physical/mental/overlap) in both age groups and for all diagnosis/symptom complexes in adults and most diagnosis/symptom complexes in children and adolescents. These results suggest that long-term sequelae of COVID-19 may play a role in both younger and older age groups.

Our study is in line with previous epidemiological studies among adults including a control group of persons not previously exposed to SARS-CoV-2 infections that observed about twofold higher odds of persisting health symptoms [8,9,19]. Other studies based on large health insurance data sets covering inpatient as well as outpatient health records from COVID-19 patients and matched controls have consistently reported higher health care services use and new onset of documented health conditions, including neurological, psychiatric, cardiometabolic, pulmonary, and renal complications following the acute phase of the COVID-19 disease. However, these studies have almost exclusively focused on adults [6,7,20–22]. Only one previous study that used large healthcare administrative datasets included children and adolescents [20]. The authors reported that increased post COVID-19 morbidity was recorded in children but did not present specific data or stratified analyses for this age group [20].

Our study therefore extends previous research as it is the first that suggests relevant post COVID-19 healthcare utilization and new-onset morbidity patterns documented by physicians in children and adolescents following COVID-19 disease in a large sample of patients with confirmed COVID-19 compared to a matched control group. Previous studies among children and adolescents, which did not observe significant group differences between children and adolescents with COVID-19 and controls were limited by restrictions to hospitalized patients [10], high drop-out rates and/or high risk of selection bias [10,23], self-reported outcome assessment [5,10,19,24,25], lack of a control group [10], insufficiently long follow-up time to assess post COVID-19 outcomes [23,26], and low sample size resulting in low statistical power [26,27].

The fact that our study replicates previous studies on post COVID-19 outcome patterns in adults [6,7,20,22] is an important argument for the validity of the new evidence reported for children/adolescents. Our study suggests that children and adolescents with COVID-19 disease may be at increased long-term risk for a broad spectrum of medical conditions including malaise/fatigue/exhaustion, cough, throat/chest pain, adjustment disorder, somatization disorder, headache, fever, anxiety disorder, abdominal pain, and depression. Particularly post COVID-19 mental health problems appear to be more frequent in children and adolescents relative to the control group. Adverse post COVID-19 pulmonary outcomes, however, appear to be less frequent in children and adolescents compared to adults.

These findings should be taken into account when assessing the overall risk of SARS-CoV-2 infections. Together with evidence on other relevant aspects not addressed in this study, our results may contribute to recent discussions on preventive measures, including the risk-to-benefit ratio of COVID-19 vaccinations in children and adolescents. In this regard, however, it also should be considered that incidence rates in children/adolescents with COVID-19 were generally lower than those in adults. Given similar *relative* magnitudes of post COVID-19 outcome incidence, the estimated long-term sequelae of COVID-19 therefore appear to be less pronounced in children and adolescents in *absolute* terms.

### Strengths and limitations

The main strength of our analysis is its broad database including more than 150,000 individuals with available data in the post COVID-19 phase. This unselected sample from all over Germany covers both outpatient and inpatient care and, thus, constitutes a unique and comprehensive source of evidence. The 96 outcomes considered in this study were selected based on published evidence and clinical expertise and provided a sound and sensitive basis for investigation of potential long-term sequelae of COVID-19 across multiple diagnosis/symptom complexes. Our analysis is based on documented, confirmed diagnoses made by physicians and psychotherapists. Accordingly, our results are not subject to possible distortions resulting from selective, incomplete, or inadequate self-reporting of symptoms but instead rely on information provided by medical professionals. To avoid confounding of the relationships between outcomes and exposure, we applied matching on relevant covariates, i.e. age, sex, and several prevalent medical conditions. The resulting distributions of covariates in the COVID-19 and control cohorts were similar, which indicated successful balancing. Overall, our results for adults are in accordance with those of previous, international studies based on routine health data [6,7,20,22]. This similarity suggests that external validity is high and provides indirect support for the validity of our findings for children and adolescents. Data preparation and analysis in accordance with the GPS of the DGEpi additionally supports the validity and reliability of our results.

Due to the observational nature of our study, a main limitation is that its design does not induce a causal interpretation of results. This limitation is inherent to all observational studies and, thus, virtually all studies on post COVID-19. We cannot exclude that our results may be affected by unmeasured confounding, although we minimized differences between COVID-19 and control cohort via matching. Our results may also be subject to detection bias that may arise if the health status of individuals after onset of COVID-19 was more closely monitored and better documented by physicians. Individuals with mild or asymptomatic course of COVID-19 are likely to be underrepresented in our study because SARS-CoV-2 infections may not have been documented [28], especially in the first months of the pandemic. The resulting selection of more severe COVID-19 cases may lead to higher incidence estimates in this cohort. By the same token, individuals with undocumented SARS-CoV-2 infection may have been included in the control cohort. To the extent that post COVID-19 also occurred in persons with undocumented infections, this misclassification induces an over-estimation of incidence rates in the control group and, thus, a bias towards the null in estimators of incidence rate ratios. This may particularly be a concern in the analyses of children/adolescents, as acute COVID-19 symptoms are more frequently mild and/or absent in this group so that they may not have resulted in a clinical consultation and thus not have been documented in the data used for this study. Generally, we assume that diagnoses made by physicians and psychotherapists have higher validity than self-reported outcomes. However, this may not be true for all coded diagnoses, since the use of certain diagnosis might be relevant for billing purposes. Follow-up times after COVID-19 diagnosis in our sample were limited because currently data are available up to the end of 2020 only. Accordingly, investigation of post COVID-19 in the very long run was not possible. Finally, the dynamic of the COVID-19 pandemic during the study period in 2020 was different than it is currently. Accordingly, utilization of health services has changed over time, and is not necessarily the same in 2021 as it was early in the pandemic. Hence, long-term and ongoing observation is required to assess the generalizability of our results to later phases of the pandemic.

## Conclusions

We found that COVID-19 diagnosis was associated with higher long-term demand for healthcare services as reflected in outpatient and inpatient diagnoses of a broad set of outcomes more than three months after detection of SARS-CoV-2 infection. While children and adolescents appear to be less affected than adults, these findings are statistically significant for all age groups. In children and adolescents, the incidence rate ratio of documented new-onset mental health problems during follow-up was higher compared to adults, whereas the opposite was true for documented health outcomes of the pulmonary diagnosis/symptom complex. To the extent that these results reflect a higher long-run morbidity related to SARS-CoV-2 infections, they may indicate an important public health challenge that should be considered in discussions about adequate preventive measures. Of course, these discussions must also consider harmful side effects of preventive measures [26].

In summary, our findings demonstrate that post COVID-19 cannot be dismissed among children and adolescents. Controlled population-based studies and further in-depth analyses are required to confirm results among children and adolescents and their impact for individuals and health care systems. Future analyses need to include currently unavailable, more recent data for extended follow-up of individuals with post COVID-19 condition identified in the present study and to get further insight into risk and protective factors. As more recent data become available it will also be possible to assess the persistence of the COVID-19-related documented morbidity during later phases of the pandemic.

## Supporting information

STROBE checklist

## Data Availability

Individual-level data are not publicly available due to legal and data protection restrictions. Aggregate statistics are available upon reasonable request to the authors.

## Ethics

The competent authority of the Federal State of Saxony, Germany approved the use of routine health insurance data for this study (reference number: 31-5221.40-15/68). The ethics committee of the TU Dresden approved this study (approval number: BO-EK (COVID)-482102021).

## Appendix

### Matching results

**Table A1:** Characteristics of children and adolescents in COVID-19 and control cohort after matching (full sample of 57,763 individuals with COVID-19)

| Variable | Category | n COVID-19 | Percent COVID-19 | n Control | Percent Control |
| --- | --- | --- | --- | --- | --- |
| Age | 0-11 | 31,345 | 54.3% | 156,725 | 54.3% |
|  | 12-17 | 26,418 | 45.7% | 132,090 | 45.7% |
| Sex | female | 28,120 | 48.7% | 140,600 | 48.7% |
|  | male | 29,643 | 51.3% | 148,215 | 51.3% |
| Asthma |  | 595 | 1.0% | 2,984 | 1.0% |
| Bronchopulmonary dysplasia |  | 30 | 0.1% | 125 | 0.0% |
| Cancer |  | 130 | 0.2% | 639 | 0.2% |
| Congenital heart disease |  | 868 | 1.5% | 4,303 | 1.5% |
| Diabetes: type I with insulin |  | 173 | 0.3% | 830 | 0.3% |
| Dialysis |  | 9 | 0.0% | 13 | 0.0% |
| Down syndrome |  | 70 | 0.1% | 342 | 0.1% |
| Epilepsy |  | 501 | 0.9% | 2,462 | 0.9% |
| Immunosuppressive disease |  | 474 | 0.8% | 2,354 | 0.8% |
| Immunosuppressive therapy |  | 110 | 0.2% | 534 | 0.2% |
| Obesity |  | 79 | 0.1% | 448 | 0.2% |
| Primary immunodeficiency |  | 263 | 0.5% | 1,275 | 0.4% |
| Psychomotor deficit |  | 1,577 | 2.7% | 7,919 | 2.7% |

**Table A2:**
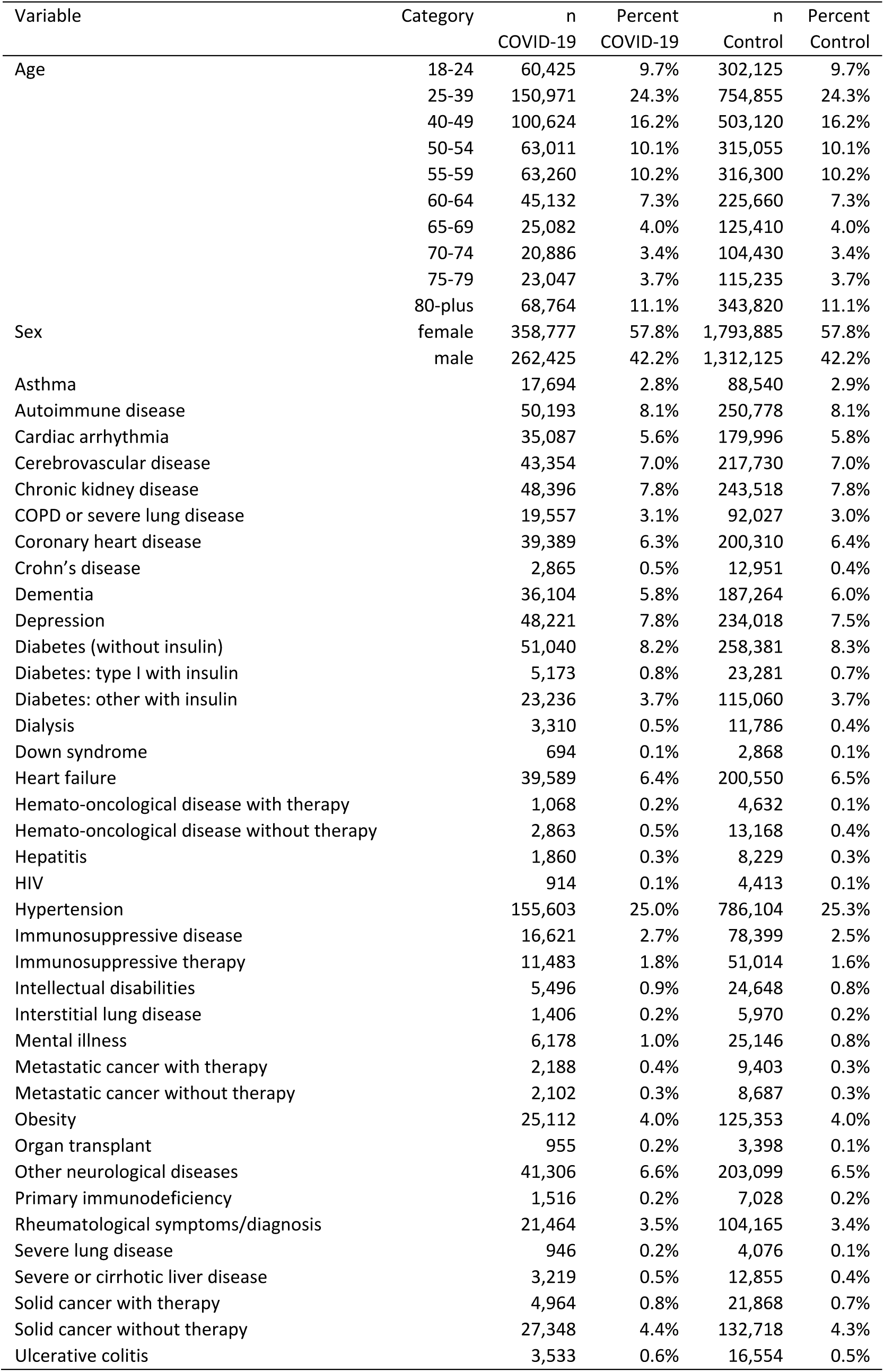
Characteristics of adults in COVID-19 and control cohort after matching (full sample of 621,202 individuals with COVID-19)

| Variable | Category | n<br>COVID-19 | Percent<br>COVID-19 | n<br>Control | Percent<br>Control |
| --- | --- | --- | --- | --- | --- |
| Age | 18-24 | 60,425 | 9.7% | 302,125 | 9.7% |
|  | 25-39 | 150,971 | 24.3% | 754,855 | 24.3% |
|  | 40-49 | 100,624 | 16.2% | 503,120 | 16.2% |
|  | 50-54 | 63,011 | 10.1% | 315,055 | 10.1% |
|  | 55-59 | 63,260 | 10.2% | 316,300 | 10.2% |
|  | 60-64 | 45,132 | 7.3% | 225,660 | 7.3% |
|  | 65-69 | 25,082 | 4.0% | 125,410 | 4.0% |
|  | 70-74 | 20,886 | 3.4% | 104,430 | 3.4% |
|  | 75-79 | 23,047 | 3.7% | 115,235 | 3.7% |
|  | 80-plus | 68,764 | 11.1% | 343,820 | 11.1% |
| Sex | female | 358,777 | 57.8% | 1,793,885 | 57.8% |
|  | male | 262,425 | 42.2% | 1,312,125 | 42.2% |
| Asthma |  | 17,694 | 2.8% | 88,540 | 2.9% |
| Autoimmune disease |  | 50,193 | 8.1% | 250,778 | 8.1% |
| Cardiac arrhythmia |  | 35,087 | 5.6% | 179,996 | 5.8% |
| Cerebrovascular disease |  | 43,354 | 7.0% | 217,730 | 7.0% |
| Chronic kidney disease |  | 48,396 | 7.8% | 243,518 | 7.8% |
| COPD or severe lung disease |  | 19,557 | 3.1% | 92,027 | 3.0% |
| Coronary heart disease |  | 39,389 | 6.3% | 200,310 | 6.4% |
| Crohn's disease |  | 2,865 | 0.5% | 12,951 | 0.4% |
| Dementia |  | 36,104 | 5.8% | 187,264 | 6.0% |
| Depression |  | 48,221 | 7.8% | 234,018 | 7.5% |
| Diabetes (without insulin) |  | 51,040 | 8.2% | 258,381 | 8.3% |
| Diabetes: type I with insulin |  | 5,173 | 0.8% | 23,281 | 0.7% |
| Diabetes: other with insulin |  | 23,236 | 3.7% | 115,060 | 3.7% |
| Dialysis |  | 3,310 | 0.5% | 11,786 | 0.4% |
| Down syndrome |  | 694 | 0.1% | 2,868 | 0.1% |
| Heart failure |  | 39,589 | 6.4% | 200,550 | 6.5% |
| Hemato-oncological disease with therapy |  | 1,068 | 0.2% | 4,632 | 0.1% |
| Hemato-oncological disease without therapy |  | 2,863 | 0.5% | 13,168 | 0.4% |
| Hepatitis |  | 1,860 | 0.3% | 8,229 | 0.3% |
| HIV |  | 914 | 0.1% | 4,413 | 0.1% |
| Hypertension |  | 155,603 | 25.0% | 786,104 | 25.3% |
| Immunosuppressive disease |  | 16,621 | 2.7% | 78,399 | 2.5% |
| Immunosuppressive therapy |  | 11,483 | 1.8% | 51,014 | 1.6% |
| Intellectual disabilities |  | 5,496 | 0.9% | 24,648 | 0.8% |
| Interstitial lung disease |  | 1,406 | 0.2% | 5,970 | 0.2% |
| Mental illness |  | 6,178 | 1.0% | 25,146 | 0.8% |
| Metastatic cancer with therapy |  | 2,188 | 0.4% | 9,403 | 0.3% |
| Metastatic cancer without therapy |  | 2,102 | 0.3% | 8,687 | 0.3% |
| Obesity |  | 25,112 | 4.0% | 125,353 | 4.0% |
| Organ transplant |  | 955 | 0.2% | 3,398 | 0.1% |
| Other neurological diseases |  | 41,306 | 6.6% | 203,099 | 6.5% |
| Primary immunodeficiency |  | 1,516 | 0.2% | 7,028 | 0.2% |
| Rheumatological symptoms/diagnosis |  | 21,464 | 3.5% | 104,165 | 3.4% |
| Severe lung disease |  | 946 | 0.2% | 4,076 | 0.1% |
| Severe or cirrhotic liver disease |  | 3,219 | 0.5% | 12,855 | 0.4% |
| Solid cancer with therapy |  | 4,964 | 0.8% | 21,868 | 0.7% |
| Solid cancer without therapy |  | 27,348 | 4.4% | 132,718 | 4.3% |
| Ulcerative colitis |  | 3,533 | 0.6% | 16,554 | 0.5% |

### Full results for domains and diagnosis/symptom complexes

**Table A3:**
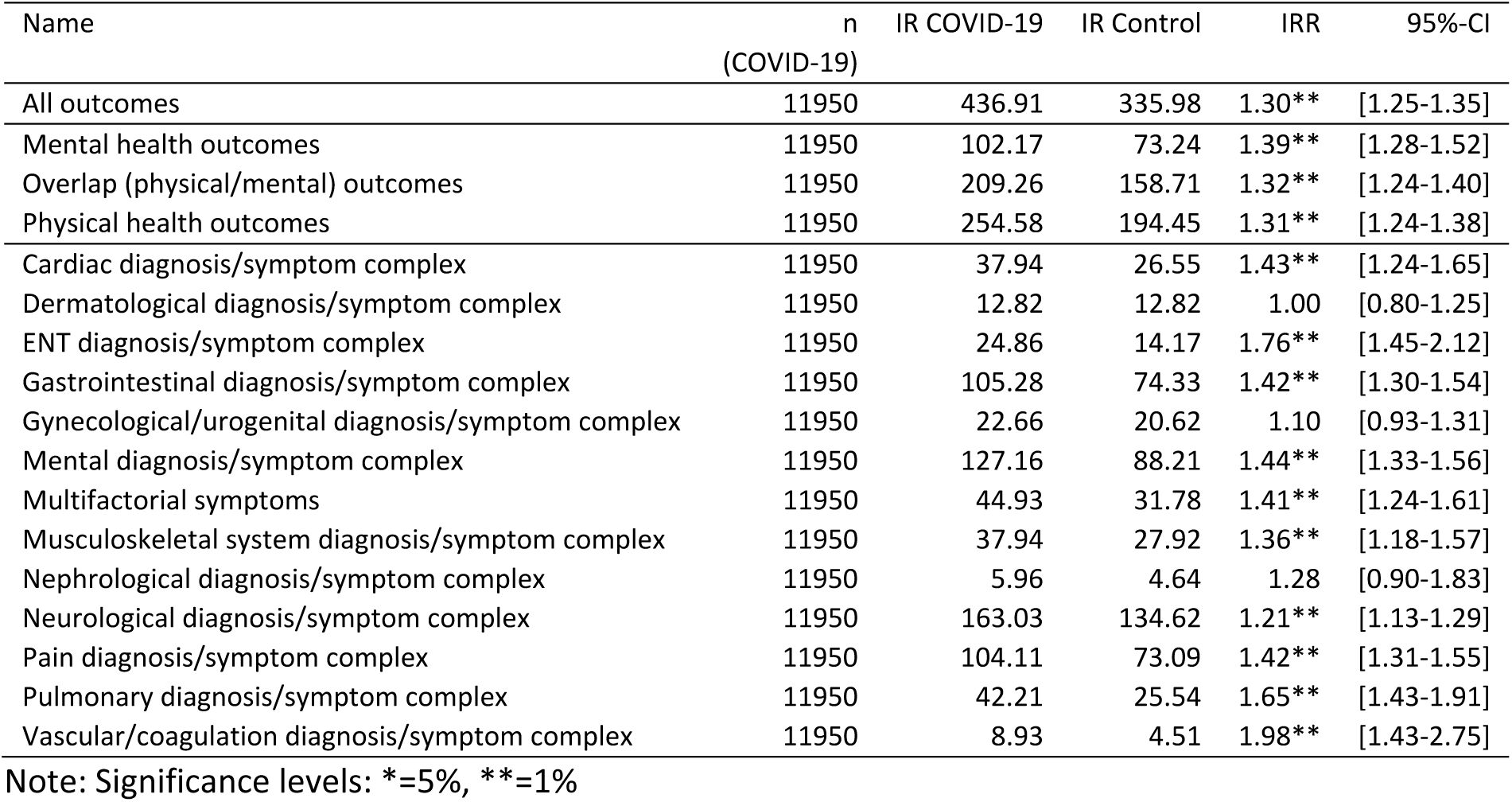
Full results for children and adolescents

**Table A4:** Full results for adults

| Name | n<br>(COVID-19) | IR COVID-19 | IR Control | IRR | 95%-CI |
| --- | --- | --- | --- | --- | --- |
| All outcomes | 145184 | 615.82 | 464.15 | 1.33** | [1.31-1.34] |
| Mental health outcomes | 145184 | 215.62 | 169.50 | 1.27** | [1.25-1.29] |
| Overlap (physical/mental) outcomes | 145184 | 278.58 | 192.59 | 1.45** | [1.42-1.47] |
| Physical health outcomes | 145184 | 422.87 | 304.42 | 1.39** | [1.37-1.41] |
| Cardiac diagnosis/symptom complex | 145184 | 144.64 | 99.20 | 1.46** | [1.43-1.49] |
| Dermatological diagnosis/symptom complex | 145184 | 23.96 | 14.40 | 1.66** | [1.58-1.76] |
| ENT diagnosis/symptom complex | 145184 | 67.05 | 42.14 | 1.59** | [1.54-1.64] |
| Gastrointestinal diagnosis/symptom complex | 145184 | 144.09 | 99.44 | 1.45** | [1.42-1.48] |
| Gynecological/urogenital diagnosis/symptom complex | 145184 | 51.41 | 46.42 | 1.11** | [1.07-1.14] |
| Mental diagnosis/symptom complex | 145184 | 275.79 | 206.30 | 1.34** | [1.32-1.36] |
| Multifactorial symptoms | 145184 | 94.74 | 69.00 | 1.37** | [1.34-1.41] |
| Musculoskeletal system diagnosis/symptom complex | 145184 | 70.81 | 54.81 | 1.29** | [1.25-1.33] |
| Nephrological diagnosis/symptom complex | 145184 | 20.69 | 14.83 | 1.39** | [1.32-1.47] |
| Neurological diagnosis/symptom complex | 145184 | 202.23 | 148.79 | 1.36** | [1.34-1.38] |
| Pain diagnosis/symptom complex | 145184 | 186.04 | 129.71 | 1.43** | [1.41-1.46] |
| Pulmonary diagnosis/symptom complex | 145183 | 83.34 | 31.81 | 2.62** | [2.53-2.71] |
| Vascular/coagulation diagnosis/symptom complex | 145184 | 71.73 | 57.73 | 1.24** | [1.21-1.28] |

Note: Significance levels: \*=5%, \*\*=1%

## References

1. Callard F, Perego E. How and why patients made long covid. Social Science & Medicine. 2021;268:113426. doi:https://doi.org/10.1016/j.socscimed.2020.113426

2. World Health Organization (WHO) clinical case definition working group on post COVID-19 condition. A clinical case definition of post COVID-19 condition by a delphi consensus. World Health Organization (WHO); Oct 2021. Available: https://www.who.int/publications/i/item/WHO-2019-nCoV-Post_COVID-19_condition-Clinical_case_definition-2021.1

3. Crook H, Raza S, Nowell J, Young M, Edison P. Long covid - mechanisms, risk factors, and management. BMJ. 2021;374:n1648. doi:10.1136/bmj.n1648

4. Augustin M, Schommers P, Stecher M, Dewald F, Gieselmann L, Gruell H, et al. Post-COVID syndrome in non-hospitalised patients with COVID-19: A longitudinal prospective cohort study. The Lancet Regional Health - Europe. 2021;6:100122. doi:https://doi.org/10.1016/j.lanepe.2021.100122

5. Sudre CH, Murray B, Varsavsky T, Graham MS, Penfold RS, Bowyer RC, et al. Attributes and predictors of long COVID. Nature Medicine. 2021;27:626–631.

6. Taquet M, Geddes JR, Husain M, Luciano S, Harrison PJ. 6-month neurological and psychiatric outcomes in 236 379 survivors of COVID-19: A retrospective cohort study using electronic health records. The Lancet Psychiatry. 2021;8:416–427. doi:https://doi.org/10.1016/S2215-0366(21)00084-5

7. Al-Aly Z, Xie Y, Bowe B. High-dimensional characterization of post-acute sequelae of COVID-19. Nature. 2021;594:259–264.

8. Akbarialiabad H, Taghrir MH, Abdollahi A, Ghahramani N, Kumar M, Paydar S, et al. Long COVID, a comprehensive systematic scoping review. Infection. 2021. doi:https://doi.org/10.1007/s15010-021-01666-x

9. Nalbandian A, Sehgal K, Gupta A, Madhavan MV, McGroder C, Stevens JS, et al. Post-acute COVID-19 syndrome. Nature Medicine. 2021;27:601–615.

10. Osmanov IM, Spiridonova E, Bobkova P, Gamirova A, Shikhaleva A, Andreeva M, et al. Risk factors for long covid in previously hospitalised children using the ISARIC Global follow-up protocol: A prospective cohort study. medRxiv. 2021. doi:10.1101/2021.04.26.21256110

11. Groff D, Sun A, Ssentongo AE, Ba DM, Parsons N, Poudel GR, et al. Short-term and Long-term Rates of Postacute Sequelae of SARS-CoV-2 Infection: A Systematic Review. JAMA Network Open. 2021;4:e2128568–e2128568. doi:10.1001/jamanetworkopen.2021.28568

12. Zimmermann P, Pittet LF, Curtis N. How common is long COVID in children and adolescents? The Pediatric Infectious Disease Journal. 2021. doi:10.1097/INF.0000000000003328

13. Michelen M, Manoharan L, Elkheir N, Cheng V, Dagens A, Hastie C, et al. Characterising long COVID: a living systematic review. BMJ Global Health. 2021;6:e005427. doi:10.1136/bmjgh-2021-005427

14. Venkatesan P. NICE guideline on long COVID. The Lancet Respiratory Medicine. 2021;9:129.

15. Hoffmann W, Latza U, Baumeister SE, Brünger M, Buttmann-Schweiger N, Hardt J, et al. Guidelines and recommendations for ensuring Good Epidemiological Practice (GEP): A guideline developed by the German Society for Epidemiology. European Journal of Epidemiology. 2019;34:301– 317.

16. Stuart EA. Matching methods for causal inference: A review and a look forward. Statistical Science. 2010;25:1–21.

17. Cameron AC, Trivedi PK. Regression analysis of count data. Cambridge: Cambridge University Press; 2013.

18. Hilbe JM. Negative binomial regression. Cambridge: Cambridge University Press; 2011.

19. Office for National Statistics. Prevalence of ongoing symptoms following coronavirus (COVID-19) infection in the UK. 2021. Available: https://www.ons.gov.uk/peoplepopulationandcommunity/healthandsocialcare/conditionsanddiseases/bulletins/prevalenceofongoingsymptomsfollowingcoronaviruscovid19infectionintheuk/1april2021

20. Taquet QAL Maxime and Dercon. Incidence, co-occurrence, and evolution of long-COVID features: A 6-month retrospective cohort study of 273,618 survivors of COVID-19. PLOS Medicine. 2021;18:1–22. doi:10.1371/journal.pmed.1003773

21. Chevinsky JR, Tao G, Lavery AM, Kukielka EA, Click ES, Malec D, et al. Late Conditions Diagnosed 1–4 Months Following an Initial Coronavirus Disease 2019 (COVID-19) Encounter: A Matched-Cohort Study Using Inpatient and Outpatient Administrative Data—United States, 1 March–30 June 2020. Clinical Infectious Diseases. 2021;73:S5–S16. doi:10.1093/cid/ciab338

22. Daugherty SE, Guo Y, Heath K, Dasmariñas MC, Jubilo KG, Samranvedhya J, et al. Risk of clinical sequelae after the acute phase of SARS-CoV-2 infection: Retrospective cohort study. BMJ. 2021;373:n1098. doi:10.1136/bmj.n1098

23. Molteni E, Sudre CH, Canas LS, Bhopal SS, Hughes RC, Antonelli M, et al. Illness duration and symptom profile in symptomatic UK school-aged children tested for SARS-CoV-2. The Lancet Child & Adolescent Health. 2021;5:708–718.

24. Miller F, Nguyen V, Navaratnam AM, Shrotri M, Kovar J, Hayward AC, et al. Prevalence of persistent symptoms in children during the COVID-19 pandemic: Evidence from a household cohort study in England and Wales. medRxiv. 2021. doi:10.1101/2021.05.28.21257602

25. Stephenson T, Pereira SP, Shafran R, De Stavola B, Rojas N, McOwat K, et al. Long COVID-the physical and mental health of children and non-hospitalised young people 3 months after SARS-CoV-2 infection; a national matched cohort study (the CLoCk) Study. Research Square. 2021. doi:https://doi.org/10.21203/rs.3.rs-798316/v1

26. Blankenburg J, Wekenborg MK, Reichert J, Kirsten C, Kahre E, Haag L, et al. Mental health of adolescents in the pandemic: Long-COVID19 or long-pandemic syndrome? medRxiv. 2021. doi:10.1101/2021.05.11.21257037

27. Radtke T, Ulyte A, Puhan MA, Kriemler S. Long-term Symptoms After SARS-CoV-2 Infection in Children and Adolescents. JAMA. 2021;326:869–871. doi:10.1001/jama.2021.11880

28. Gornyk D, Harries M, Glöckner S, Strengert M, Kerrinnes T, Bojara G, et al. SARS-CoV-2 seroprevalence in Germany - a population based sequential study in five regions. medRxiv. 2021. doi:10.1101/2021.05.04.21256597

