## Supplementary material for "Post COVID-19 in children, adolescents, and adults: results of a matched cohort study including more than 150,000 individuals with COVID-19"

### S1 Outcomes by domain and diagnosis/symptom complex

#### Outcomes by domain

**Mental health outcomes**: Adjustment disorder; Anxiety disorder; Behavioral symptoms; Cognitive function impairment; Concentration impairment/Concentration deficit; Depression; Disorientation; Emotional and behavioral disorder; Mood disorder; Obsessive-compulsive disorder; Somatization disorder

**Overlap (physical/mental) outcomes**: Abdominal pain; Acute pain; Cachexia; Changes in bowel habits; Chronic fatigue syndrome; Developmental delay; Dysuria; Eye pain; General symptoms; Headache; Hyperhidrosis; Joint pain; Loss of appetite, weight gain/loss, eating disorders; Malaise/fatigue/exhaustion; Memory impairment; Myalgia; Neurasthenia; Other coordination disorders/ataxia; Pain, not elsewhere classified; Paresthesia of skin; Post-COVID; Sensation and perception disorder; Sleep disorders; Somnolence, sopor, coma; Throat/chest pain

**Physical health outcomes**: Anuria, oliguria; Arthritides; Ascites; Carditis due to viruses; Cough; Covid toe; Diarrhea; Dysgeusia; Dyslexia; Dysmenorrhea; Dysphagia; Dyspnea; Epistaxis; Facial nerve paralysis; Fever; Flatulence; Gangraen; Hair loss; Hearing loss/tinnitus; Heart failure; Heart murmurs; Heartburn; Hemorrhage; Hepatomegaly and splenomegaly; Hoarseness; Hypotension; Impaired balance; Lymphadenopathy; Meningismus; Movement disorders; Multisystemic inflammatory syndrome; Myocardial infarction; Myocarditis; Nausea; Neurological manifestation of Post-COVID; Oedema; Other cardiac arrhythmias; Other symptoms of the urinary system; Paresis; Pathological findings from male genital tract; Pathological lung findings; Pathological reflexes; Pericarditis; Polyuria; Pulmonary embolism; Rash; Respiratory insufficiency; Seizures; Shock; Sinus vein thrombosis; Speech and language disorders; Stroke; Subcutaneous nodules; Syncope; Tachycardia/Palpitation; Tetany; Thrombosis; Urethral discharge; Urinary retention; Vertigo; Visual disturbances

### Outcomes by diagnosis/symptom complex

**Cardiac diagnosis/symptom complex**: Carditis due to viruses; Heart failure; Heart murmurs; Hypotension; Myocardial infarction; Myocarditis; Other cardiac arrhythmias; Pericarditis; Shock; Syncope; Tachycardia/Palpitation; Throat/chest pain

**Dermatological diagnosis/symptom complex**: Hair loss; Rash; Subcutaneous nodules

**ENT diagnosis/symptom complex**: Dysgeusia; Dysphagia; Epistaxis; Hearing loss/tinnitus; Hoarseness; Vertigo

**Gastrointestinal diagnosis/symptom complex**: Abdominal pain; Ascites; Changes in bowel habits; Diarrhea; Dysphagia; Flatulence; Heartburn; Hepatomegaly and splenomegaly; Nausea

**Gynecological/urogenital diagnosis/symptom complex**: Dysmenorrhea; Pathological findings from male genital tract; Urethral discharge

**Mental diagnosis/symptom complex**: Adjustment disorder; Anxiety disorder; Behavioral symptoms; Chronic fatigue syndrome; Cognitive function impairment; Concentration impairment/Concentration deficit; Depression; Disorientation; Emotional and behavioral disorder; Malaise/fatigue/exhaustion; Memory impairment; Mood disorder; Neurasthenia; Obsessive-compulsive disorder; Other coordination disorders/ataxia; Paresthesia of skin; Sensation and perception disorder; Sleep disorders; Somatization disorder; Somnolence, sopor, coma

**Multifactorial symptoms**: Acute pain; Cachexia; Fever; General symptoms; Hyperhidrosis; Loss of appetite, weight gain/loss, eating disorders; Lymphadenopathy; Oedema; Pain, not elsewhere classified

**Musculoskeletal system diagnosis/symptom complex**: Arthritides; Impaired balance; Joint pain; Movement disorders; Myalgia

**Nephrological diagnosis/symptom complex**: Anuria, oliguria; Dysuria; Other symptoms of the urinary system; Polyuria; Urinary retention

**Neurological diagnosis/symptom complex**: Chronic fatigue syndrome; Developmental delay; Dysgeusia; Dyslexia; Dysphagia; Facial nerve paralysis; Impaired balance; Meningismus; Movement disorders; Neurasthenia; Neurological manifestation of Post-COVID; Other coordination disorders/ataxia; Paresis; Paresthesia of skin; Pathological reflexes; Seizures; Sensation and perception disorder; Sinus vein thrombosis; Sleep disorders; Somnolence, sopor, coma; Speech and language disorders; Stroke; Tetany; Urinary retention; Vertigo; Visual disturbances

**Pain diagnosis/symptom complex**: Abdominal pain; Acute pain; Dysuria; Eye pain; Headache; Joint pain; Myalgia; Pain, not elsewhere classified; Throat/chest pain

**Pulmonary diagnosis/symptom complex**: Cough; Dyspnea; Pathological lung findings; Pulmonary embolism; Respiratory insufficiency

**Vascular/coagulation diagnosis/symptom complex**: Covid toe; Epistaxis; Gangraen; Hemorrhage; Myocardial infarction; Pulmonary embolism; Sinus vein thrombosis; Stroke; Thrombosis

### S2 Covariates

#### Covariates for children and adolescents

Asthma; Bronchopulmonary dysplasia; Cancer; Congenital heart disease; Diabetes: type I with insulin; Dialysis; Down syndrome; Epilepsy; Immunosuppressive disease; Immunosuppressive therapy; Obesity; Primary immunodeficiency; Psychomotor deficit

#### Covariates for adults

Asthma; Autoimmune disease; Cardiac arrhythmia; Cerebrovascular disease; Chronic kidney disease; COPD or severe lung disease; Coronary heart disease; Crohn's disease; Dementia; Depression; Diabetes (without insulin); Diabetes: type I with insulin; Diabetes: other with insulin; Dialysis; Down syndrome; Heart failure; Hemato-oncological disease with therapy; Hemato-oncological disease without therapy; Hepatitis; HIV; Hypertension; Immunosuppressive disease; Immunosuppressive therapy; Intellectual disabilities; Interstitial lung disease; Mental illness; Metastatic cancer with therapy; Metastatic cancer without therapy; Obesity; Organ transplant; Other neurological diseases; Primary immunodeficiency; Rheumatological symptoms/diagnosis; Severe lung disease; Severe or cirrhotic liver disease; Solid cancer with therapy; Solid cancer without therapy; Ulcerative colitis

### S3 Results for age groups 0-11 and 12-17

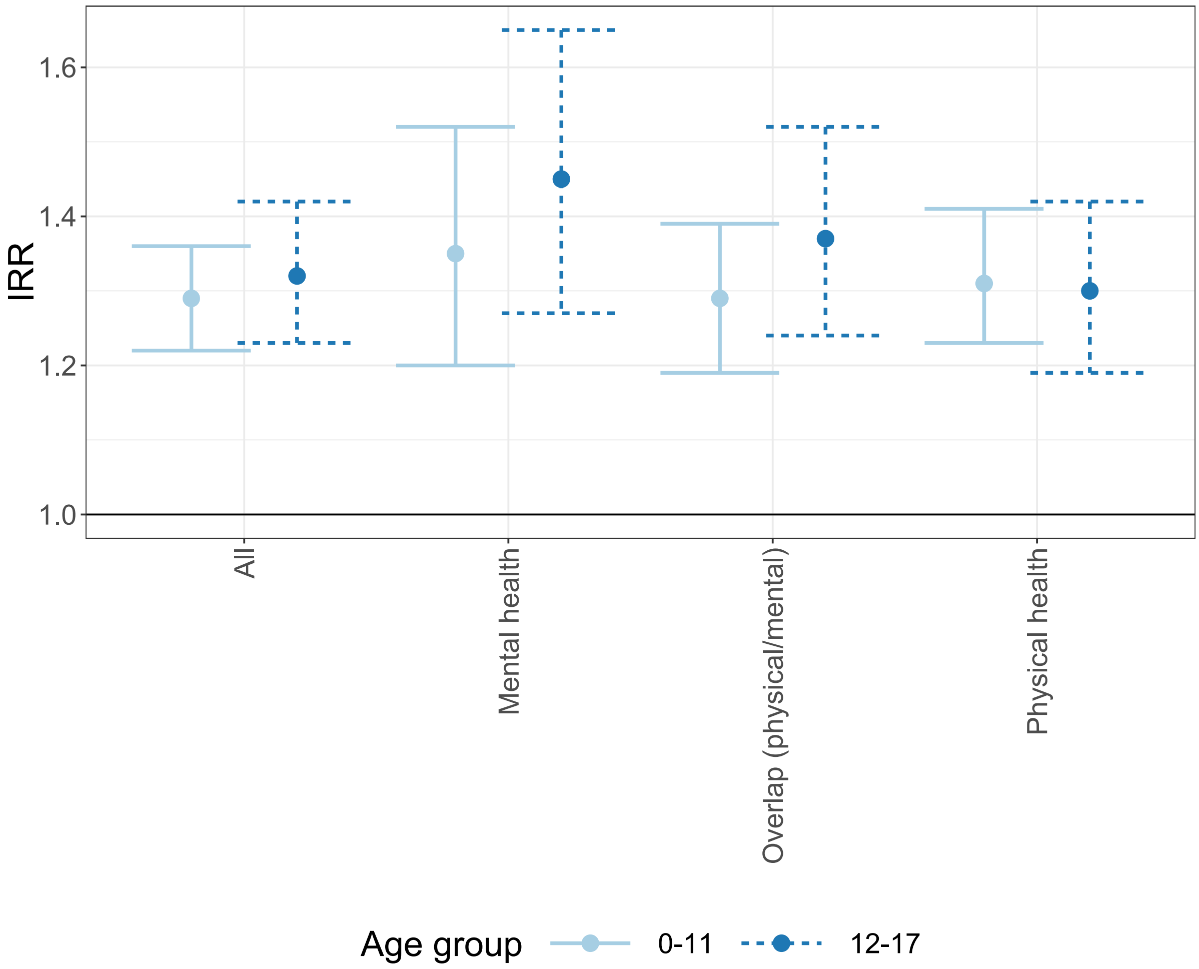

Figure S1: Estimated incidence rate ratios with 95%-confidence intervals in age groups 0-11 and 12-17 by domain

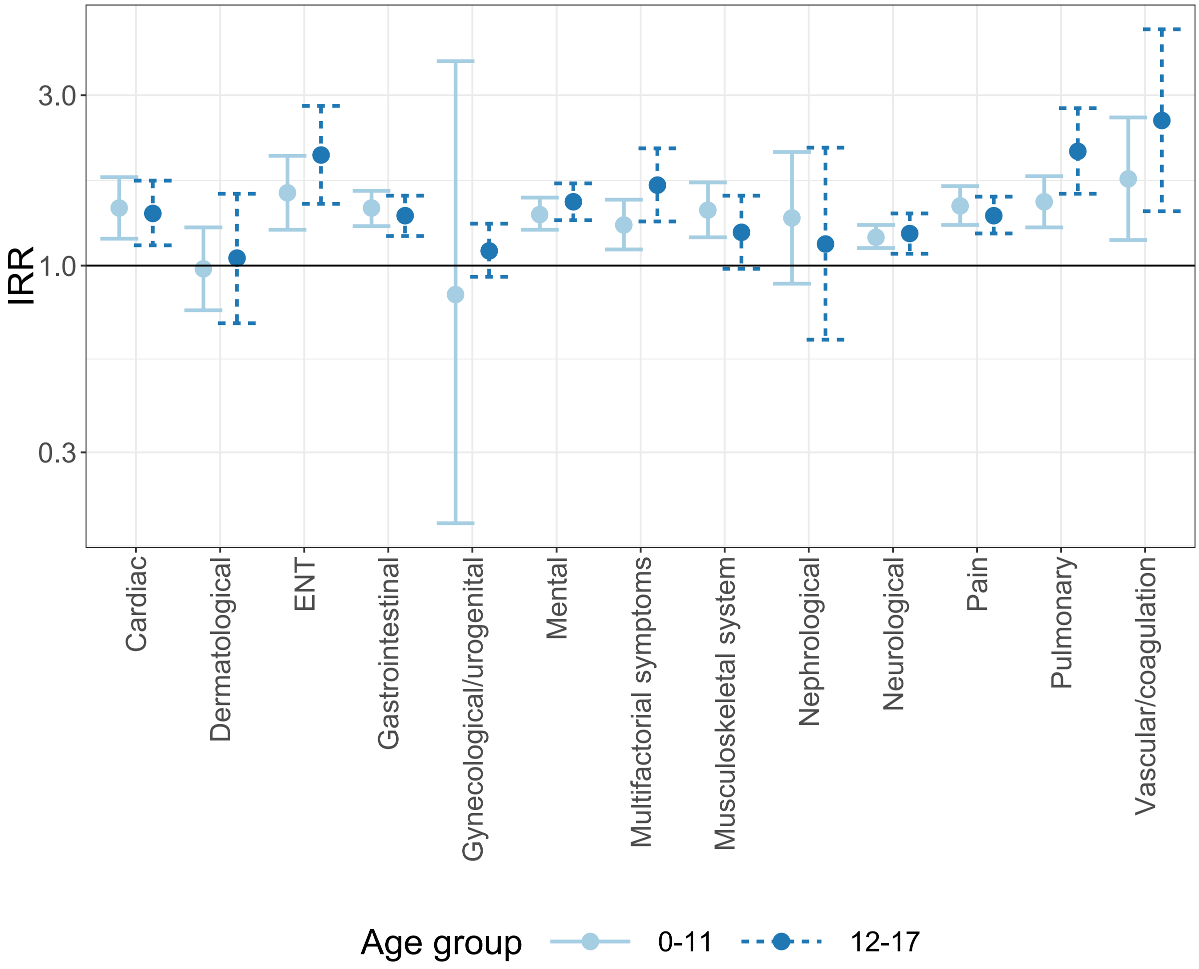


Figure S2: Estimated incidence rate ratios with 95%-confidence intervals in age groups 0-11 and 12-17 by diagnosis/symptom complex
